# Investigating the effect of respiratory indices to predict mortality and the status of trauma patients using artificial neural networks

**DOI:** 10.1101/2020.06.30.20143784

**Authors:** Zahra Hafezi, Mohammad Sabouri, Milad Shayan, Shahram Paydar

## Abstract

**Introduction:** Knowing the final status of trauma patients and clearly understanding their condition are always significant due to the complexity of injuries and the high dependence of the patient’s condition on various factors. That is why it allows doctors to be able to provide required facilities in a broader perspective and perform appropriate action. In addition, it can avoid wasting time and energy and then increasing patient mortality. While, there are several ways to measure and predict the final status of patients but all of them have some defects. Therefore, it is very important for the significance of the system design with high accuracy and reliability to be able to help physicians investigate the final status of trauma patients.

**Method:** In this study, a method, a sub-branch of data science and artificial intelligence, is presented and studied based on artificial neural networks to estimate the final status of trauma patients and predict their survival and death probabilities during the treatment and care process. In the proposed method, the final status of patients is predicted using 13 respiratory indices. This method is run in Matlab and its efficiency is studied.

**Results:** Research subjects include 3073 patients, 494 females and 2579 males, from Shahid Rajaei Medical Center in Shiraz. In general, according to the results from testing the method, it has been able to accurately predict the mortality of patients based on respiratory indices. The proposed structure has been able to predict patients’ survival and death probabilities with an accuracy of %73.75 and %99.71 respectively. Therefore, we can conclude that the presented and examined method can make a significant relevance between calculated respiratory indices and final status of patients.

**Discussion and Conclusion:** Due to the present study and the obtained results and investigating the mortality relevance with the other 13 respiratory indices using artificial intelligence-based methods, it can be stated that these indicators are good criteria for predicting mortality.

## 1. Introduction

Trauma is known as one of the main causes of mortality in the world. Therefore, it is considered important due to the researches in this field and a decrease in mortality among patients. Based on the type of injury or damage as well as the environmental conditions of incident location, traumatic patients suffer from injuries through different mechanisms depending on the type of injury and the stressful conditions of trauma, different responses are obtained from patients. In such a way that after completing the treatment period, some patients are released without specific problem. Nevertheless, passing these difficult conditions in some patients leads to their death [1]. Therefore, predicting the final status of patients will cause to improve the ability of distinguishing between survived and died patients [2]; so, such a system can be an effective way to manage patients. Therefore, knowing the status of traumatic patients and anticipating the upcoming problems has always been considered by human to be able to greatly confront different conditions ahead. Trauma surgeons are no exception. They always try to find methods to determine the status and prognosis of patients in order to be more prepared to manage what has happened and will happen to the patient. Many efforts have been made to achieve this and numerous indicators have been identified as predictive and determinant factors for prognosis. However, predicting in the early phases is one of the major problems [3] but different models for prediction have been studied so far. Many of these models used physiological and anatomical information to predict the consequences of trauma including Glasgow Coma Scale (GCS) [4], Injury Severity Scale (ISS) [5-7], New ISS [8] and Revised Trauma Score (RTS) [10]. Laboratory data have also been used in these models [1]. Due to the frequency of multiple trauma injuries and the importance of its treatment and rehabilitation in the shortest time and determine the prognosis of patients, various studies have been performed on the factors predicting the casualty’s malaise. Ouellet et al. [10] investigated a study of 2269 patients examined the value of blood gas analysis in predicting mortality in trauma patients and found that there is a significant relevance between blood gasses, particularly lactate and BD levels and mortality and length of stay in a hospital. In a similar study, however, Gale et al. [11] compared the value of lactate serum level and open deficiency in mortality prediction of trauma patients. They evaluated 1,829 patients and found that both lactate and BD levels in died persons have been reported to be very high and both of them predict premature mortality. Reed et al. [12] designed a system called Respiratory Index of Severity in Children (RISC) in order to predict the death from lower respiratory tract disease in children and stated that their system makes a good distinction between children with lower respiratory tract disease. It is also a tool to determine the severity of respiratory diseases in children based on mortality risk. Following this, Emukule et al. [13] presented a modified type of RISC called mRISC (modified Respiratory Index of Severity in Children) and there were able to specify children who are at risk of death due to respiratory system diseases.

According to previous researches and studies on the factors determining and measuring factors conducted to assess the prognosis of trauma patients as well as the high relevance of respiratory indicators for mortality prediction among traumatic patients, we can consider them as good criteria for the rapid diagnosis of tissue hemorrhagic conditions in trauma patients [10] and determining patient prognosis [14].

Due to the high ability of the respiratory system to regulate blood pressure, the type of blood gasses’ changes associated with the number of breaths can indicate a function level of the respiratory system. It can be also used as a predictive factor for the patient’s respiratory condition within a few vital hours after trauma. By investigating previous researches to determine the factors that predict the assessment of the patients’ status, it is expected that investigation and analysis of respiratory indicators in this field can determine the prognosis of casualty.

Neural networks have been used for five decades to analyze medical data and because of the ability to learn on complex issues as well as maintain an accuracy, they have always considered even in analysis of some data [15,16], including death prediction using statistical methods [11]. Among the successful studies conducted by these artificial networks, we can point out the studies of Soni et al. who used the neural network to predict heart disease[17]. They used the data collected by Rajkumar [18]. Wang et al. also conducted a study on 8640 patients to diagnose type 2 diabetes. They performed their experiment with an artificial neural network and a multivariate logistics network. They compared the results and concluded that the accuracy of the artificial network is higher [19]. Wajs et al. could have predicted arterial blood gas by using neural and arterial gas data from experiments repeated over the specific periods of time [20].

In this study, different respiratory indices have been used and assessed in new formulas to determine whether these indicators can predict patient prognosis and mortality: Respiratory index (RI) or the ratio of arterial alveolar oxygen gradient (A-a oxygen gradient). Arterial oxygen is a measure of the difference in oxygen concentration between the alveoli in lungs and arterial blood and its changes indicate them in the body’s oxygenation. RI can be easily calculated by the values in the blood gas test, and the normal values vary according to the individuals’ age that is increasing with age [21]. RI is an indicator increasing in some diseases such as liver, heart and sepsis [22] and especially lung diseases [22, 23]. Therefore, this index has been widely used to evaluate lung function and severity of lung injuries [23, 24].

Previous studies have tried to use alveolar-arterial oxygen gradient as a diagnostic method for pulmonary embolism [21]. Therefore, a normal index, in other words, normal alveolar-arterial oxygen gradient in a patient with no history of pulmonary embolism or deep vein thrombosis significantly reduces the chances of pulmonary embolism. If doctors are less likely to have an embolism in the initial examination, measurement of alveolar-arterial oxygen gradient can provide sufficient evidence to refuse the diagnosis [21]. However, this has not been confirmed for future studies in pregnant patients. As mentioned, alveolar-arterial oxygen gradient cannot be used as a pulmonary embolism screening method in pregnant patients. So, imaging procedures should be performed for these patients as soon as possible [15].

Another study is aimed to examine RI as a factor expressing severity in community-acquired pneumonia [16]. According to this study, it was shown that decreased blood oxygen levels led to increased respiratory index are associated with myocardial damage and an increase in troponin. This can be used as a method to determine the severity of pneumonia.

This paper presents an approach based on artificial neural network to investigate the relevance between respiratory indices and mortality among trauma patients and its efficiency and, in fact, it is a continuation of studies in the field of arterial blood gases [25, 26]. As can be seen in these two studies[25, 26], artificial intelligence methods are very effective in predicting medical parameters. This paper is organized as follows. Section 2 presents the basic concepts of the proposed method and the nature of the used data. The results of the performed simulation is presented in Section 3. Finally, section 4 discusses and concludes the results of present study.

## 2. Methods

The purpose of present study is to find a relevance between the types of respiratory and mortality indices and a method for predicting mortality and the final status of patients conducted in the trauma research department of Shahid Rajaei hospital, Shiraz, based on data in 2016-2017. In this study, 3915 subjects participated and 494 females and 2579 males were selected with complete data. Respiratory indices are calculated for them indicated in Table 1. Table 1 shows the formulas experimentally obtained and studied in Shahid Rajaei hospital in Shiraz. This study is investigated the relevance among these 13 indexes based on mortality.

**Table 1.**
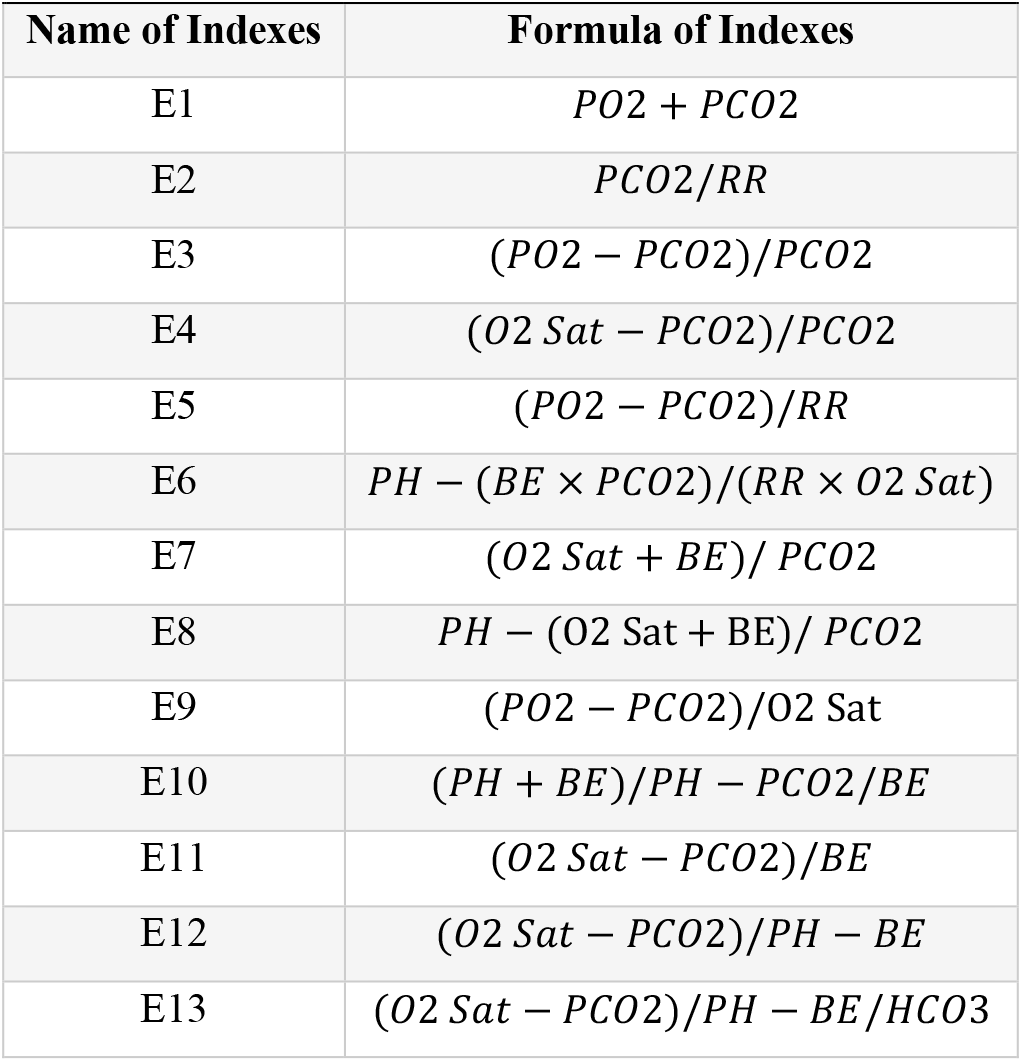
Respiratory indices

| Name of Indexes | Formula of Indexes |
| --- | --- |
| E1 | $PO_2 + PCO_2$ |
| E2 | $PCO_2/RR$ |
| E3 | $(PO_2 - PCO_2)/PCO_2$ |
| E4 | $(O_2 Sat - PCO_2)/PCO_2$ |
| E5 | $(PO_2 - PCO_2)/RR$ |
| E6 | $PH - (BE \times PCO_2)/(RR \times O_2 Sat)$ |
| E7 | $(O_2 Sat + BE)/PCO_2$ |
| E8 | $PH - (O_2 Sat + BE)/PCO_2$ |
| E9 | $(PO_2 - PCO_2)/O_2 Sat$ |
| E10 | $(PH + BE)/PH - PCO_2/BE$ |
| E11 | $(O_2 Sat - PCO_2)/BE$ |
| E12 | $(O_2 Sat - PCO_2)/PH - BE$ |
| E13 | $(O_2 Sat - PCO_2)/PH - BE/HCO_3$ |

### Neural Network Topology

The proposed method is based on finding the relevance between the calculated and mortality indices and using a topology based on artificial neural networks. This consists of two artificial neural networks. One of them analyzes the data and finds a relevance between input data and died patients. Another one finds a relevance with the status of alive patients. The proposed structure is composed of a combination of two networks and a comparator shown in Figure 1.

**Figure 1.**
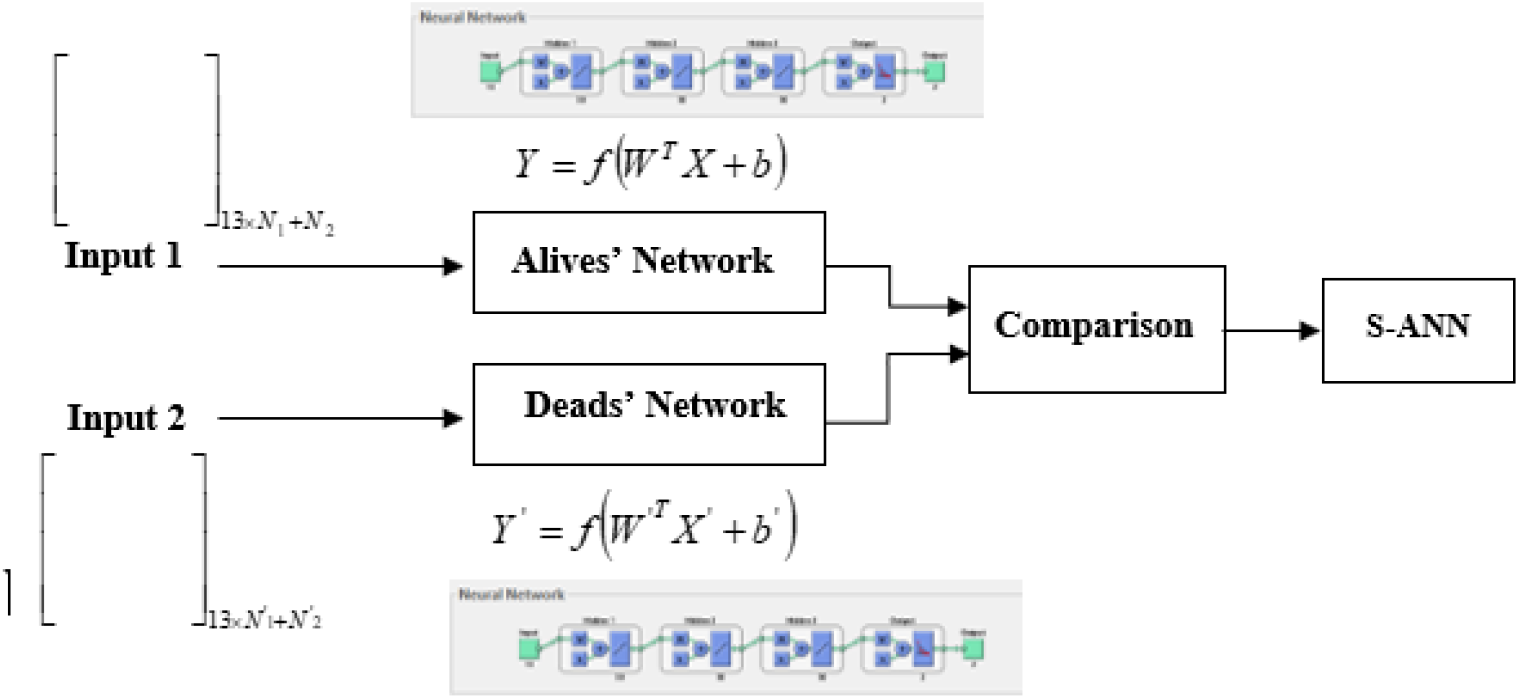
The general structure of the neural network used in this study

Both Deads and Alives networks have a topology of multilayer perceptron (MLP) in three hidden layers and 50 neurons in each layer. These properties have been selected based on several tests and experiments. Table 2 shows some of the performed tests to find the best network structure. In the process of tests performed for each of the networks described in Table 2, the percentage of network training and testing accuracy was calculated and the best network with the highest output percentage was selected.

**Table 2.** Accuracy of training and test of selected neural networks

| Network Name | Number of hidden layers | Number of Neurons in Hidden layer 1 | Number of Neurons in Hidden layer 2 | Number of Neurons in Hidden layer 3 | Number of Neurons in Hidden layer 4 |
| --- | --- | --- | --- | --- | --- |
| **Network 1** | 2 | 20 | 20 | --- | --- |
| **Network 2** | 3 | 20 | 20 | 20 | --- |
| **Network 3** | 3 | 50 | 50 | 50 | --- |
| **Network 4** | 3 | 80 | 80 | 80 | --- |
| **Network 5** | 4 | 20 | 20 | 20 | 20 |
| **Network 6** | 4 | 80 | 80 | 80 | 80 |

### Evaluation

In order to measure the accuracy of the proposed structure and its performance after modeling and training of neural networks, the results should be evaluated. In order to evaluate the new data set, 687 alive and 213 dead samples were used in A-ANN and output results compared to real samples to show network efficiency. Among them, Mean Relative Error (MRE) and Mean Square Error (MSE) are the most common and used measures in this study based on the following equations:

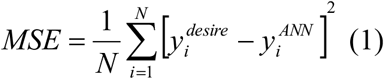

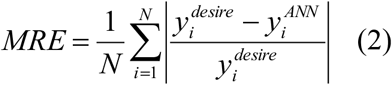

## 3. Results

The present study was involved 3,915 patients, but the data of 3037 patients were applied to study. In such a way that 2322 and 751 patients were used directly (education process) and indirectly (verification process) in an average age of 37.2, respectively. ABG and VS values were recorded for all patients in early hospital arrival. Therefore, based on these values, 13 indices (raw data) were calculated based on the formulas listed in Table 1.

The following results were obtained by training the structure presented in Figure 1 in different modes and structural properties in the networks and each one is finally verified. Table 3 presents the results of each network test. In addition, it is obvious that Network 3 has been able to perform the best function in 76% accuracy among the other networks.

**Table 3.** Accuracy of training and test of selected neural networks

| Network Name | Alives Accuracy(%) | Deads Accuracy (%) | A-ANN Accuracy (%) |
| --- | --- | --- | --- |
| **Network 1** | 99.41% | 96.87% | 71.72% |
| **Network2** | 99.41% | 96.87% | 72.58% |
| **Network 3** | **99.71%** | **93.75%** | **76.00%** |
| **Network 4** | 99.85% | 95.31% | 72.18% |
| **Network 5** | 98.98% | 100% | 71.02% |
| **Network 6** | 99.41% | 95.31% | 69.44% |

Table 4 separately indicates the results from training and testing of selected network and proposed structure. In addition, according to this table and equations 1 and 2 for A-ANN network, two values of final MSE and final MRE have been obtained 0.2085 and 0.2543, respectively, showing the appropriate efficiency of the network.

**Table 4.**
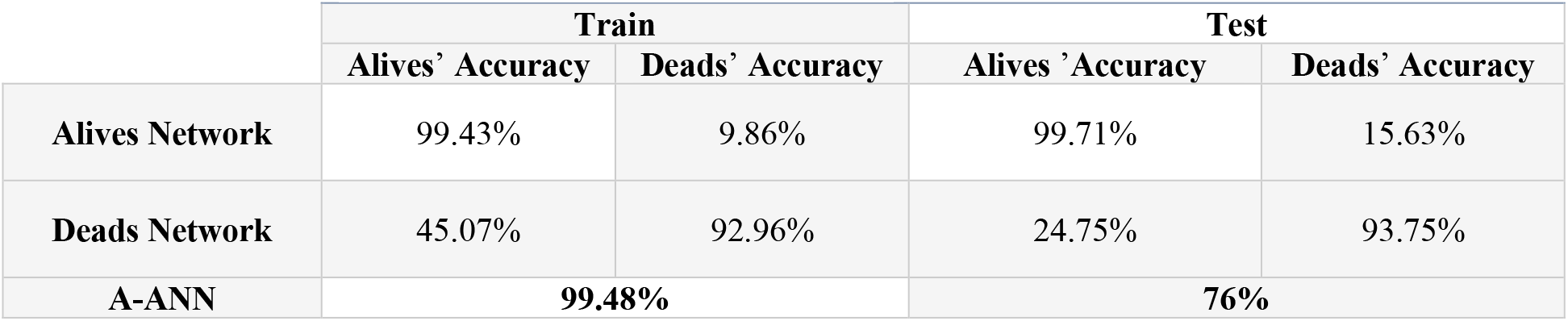
Train and Test Accuracy

|  | Train |  | Test |  |
| --- | --- | --- | --- | --- |
|  | Alives' Accuracy | Deads' Accuracy | Alives' Accuracy | Deads' Accuracy |
| **Alives Network** | 99.43% | 9.86% | 99.71% | 15.63% |
| **Deads Network** | 45.07% | 92.96% | 24.75% | 93.75% |
| **A-ANN** | **99.48%** |  | **76%** |  |

Figures 2 and 3 show the output and results of the training and testing process of dead and alive networks with training and testing data.

**Figure 2.**
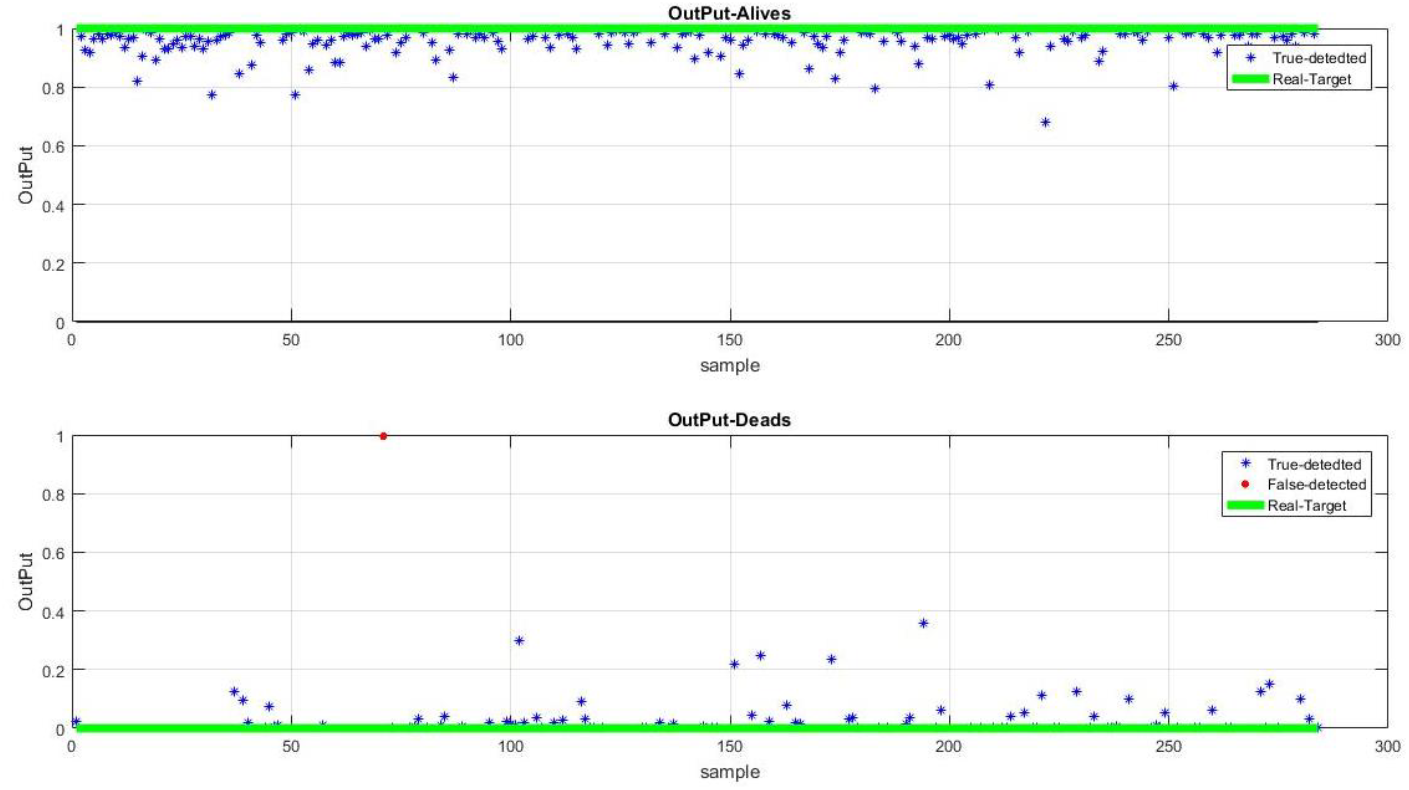
A-ANN outputs for Alives and Deaths sample in training dataset

**Figure 3.**
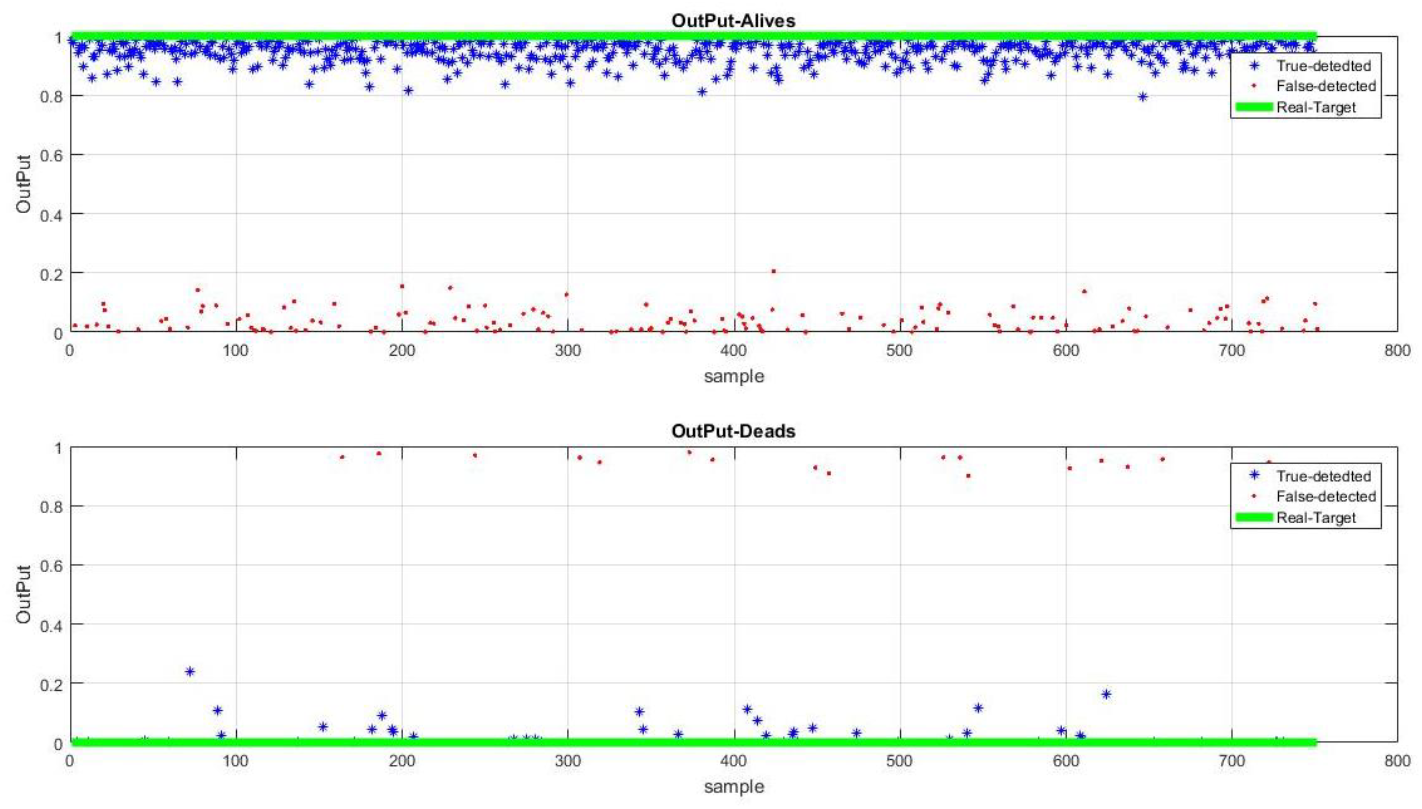
A-ANN outputs for Alives and Deaths sample in testing dataset

According to presented results, the proposed method has been able to accurately calculate about 76% mortality using respiratory indices and find a significant relevance between these indices and output.

## 4. Discussion and Conclusion

This study is purposed to investigate the data relating to 3073 severe trauma patients referred to resuscitation room of Rajaei hospital, Iran. This dataset based on 277 deaths and 2796 lives was analyzed to find a way to predict mortality of trauma patients based on different respiratory indices and formulas. However, some broad studies have been performed on the patients’ prognosis; in this case, several methods have been studied and analyzed. However, researchers have always considered new methods to predict and examine the condition of patients. For example, GCS is one of the first criteria for standardizing the assessment of consciousness level of patients with brain injury presented in 1970 [27, 28]. Given the importance of this criterion and its high performance, researchers are also looking for the applied ways and methods to predict mortality. In the meantime, considering the respiratory system and its related parameters due to the high significance and its important role, especially in patients with trauma, many researchers have studied this system to find the relevance between its indicators and mortality and predict this parameter. Among the important studies, we can consider the researches of Ouellet et al. [10] on arterial blood gas and its relevance to lactate and mortality. Gale et al.[11] also found that a high lactate level has been reported among dead trauma patients. Unlike the researches of some researchers such as McFarlane [21], Deutsch [15], and Chieregato [3], who could not make a definite relevance between respiratory system behavior and mortality. However, similar studies by Reed et al. (2012) [12], Emukule [13] could be found a relevance between respiratory system and mortality. They stated that the study on the respiratory system could be extremely effective in predicting mortality. Accordingly, the present study examines the respiratory system and the indicators related to this system. Therefore, 13 formulas of different respiratory indices were examined using a method based on artificial neural networks and ROC curve. So according to the obtained results, there was a significant relevance between these indices and mortality, then, mortality prediction. This shows that the respiratory indices such as PO2, PCO2, RR, PaO2, and O2 Sat have about 76% mortality predictive power.

## Acknowledgments

The authors would like to thank the members of Shiraz Trauma Research Center of Shahid Rajaee (Emtiaz) Hospital members, especially Dr. Shahram Bolandparvaz, head of Trauma Research Center, for providing this research opportunity, and also Dr. HamidReza Abbasi, deputy of Trauma Research Center for his cooperation and supporting in this research.

## Authors’ contributions

ZH and MS and MSH contributed to the study conception, implementation of the study, design, interpretation and critical was involved in drafting of the manuscript. SHP contributed to the conception and design of data and revising the manuscript. All authors have read and approved the manuscript, and ensure that this is the case.

## Availability of data and materials

The data that support the findings of the present study are available from the Rajaee (Emtiaz) Hospital and are not publicly available. The anonymized dataset used for the present research is however available from the corresponding author on reasonable request and with permission of both Trauma Research Center and Rajaee (Emtiaz) Hospital of Shiraz.

## Ethics approval and consent to participate

This study was conducted according to the principles expressed in the Trauma Research Center of Shahid Rajaee (Emtiaz) Hospital of Shiraz and approved by the local ethics committee of Shiraz University of Medical Sciences by the code IR.SUMS.REC.1397.719. Based on the approval of Ethics Committee, all informations were collected only by patient’s code and their identity was not disclosed. Patient’s information was in private.

